# The effect of geographical variation in income measures on Measles-Mumps-Rubella uptake and coverage in England; a protocol for an ecological study

**DOI:** 10.1101/2022.12.21.22283782

**Authors:** Salma Ghazal, Ireneous N. Soyiri

## Abstract

Measles is a vaccine-preventable disease whose vaccine was introduced in the United Kingdom in 1988, however, Measles outbreaks are still occurring in England. Consequently, the World Health Organization (WHO) removed the UK’s elimination status of Measles in 2019.

Noticeably, MMR vaccination coverage in England is below the recommended threshold with geographical variations across local authorities (LA). The research into the effect of income disparities on MMR vaccine coverage was insufficiently examined. Therefore, an ecological study will be conducted aiming at determining whether there is a relationship between income deprivation measures and MMR vaccine coverage in upper-tier local authorities in England, using 2019 publicly available data. The effect of spatial clustering of income level on vaccination coverage will also be assessed.

Vaccination coverage data will be obtained from “Cover of Vaccination Evaluated Rapidly (COVER)”. Income deprivation score, Deprivation gap, and Income Deprivation Affecting Children Index will be obtained from Office for National Statistics and Moran’s Index will be generated using RStudio.

Rural/urban LA classification and mothers’ education will be included as possible confounding factors. Additionally, the live births rate per mothers’ age group will be included as a proxy for the mothers’ age variation in different LA. Multiple linear regression will be used after testing the relevant assumptions, using SPSS software. Moran’s I together with income deprivation score will be analysed through regression and mediation analysis.

This study will help in determining whether income level is a determinant of MMR vaccination uptake and coverage in LA in England which would help policymakers in designing targeted campaigns, thus preventing measles outbreaks in the future.

## 4. BACKGROUND AND RATIONALE

The use of Measles, Mumps and Rubella (MMR) trivalent vaccine began in the United Kingdom in 1988(1). However, after more than thirty years of its introduction, measles is not considered something of the past as yet, as several measles outbreaks have been occurring in England during the last decade,(2-4) which led to the UK’s Measles and Rubella’s elimination status to be removed by the World Health Organization (WHO) in 2019(4). In addition to the outbreaks, the overall vaccination coverage of measles in England is below the recommended herd immunity threshold of 95%, with noticeable geographical variations in coverage across different local authorities(5, 6).

Some studies found a relationship between the decreased uptake of MMR and low household income or unemployment(7-9). Moreover, mothers’ mental health was also found to be associated with low uptake,(10) as well as, mothers’ education level; age when she gave birth to the child; and employment status (11). Other reported factors that withheld parents from giving their children the vaccine were diverse (12). Some of them were related to access difficulties faced by some minority groups,(13) but other parents had concerns surrounding the safety of the vaccine(11, 14). A Directed Acyclic Graph (DAG) summarising the factors that were found to be associated with MMR uptake is shown in **Fig. 1** below.

**Fig. 1:**
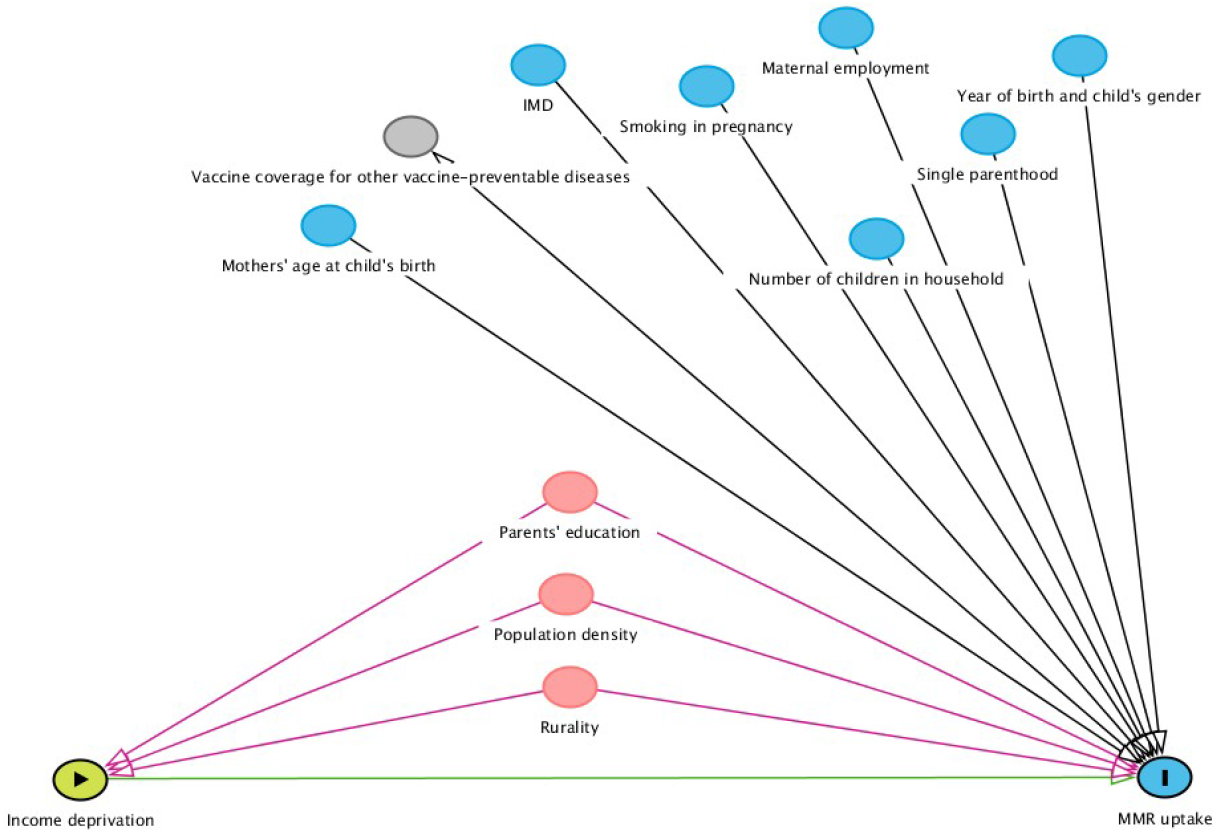
Represents a Directed Acyclic Graph (DAG) showing the factors that were found to be affecting MMR uptake in England (IMD is the Index of Multiple Deprivation). DAG was constructed using Ferguson *et al*, (2020) proposed methods for creating DAGs(19). The image was generated using DAGitty.net online software (20)

Generally, Social Determinants of Health (SDH) have been found to affect health implicitly(15) and some of them were found to affect vaccination uptake and coverage(9, 16-18). However, the association between deprivation and uptake of the MMR vaccine specifically was insufficiently examined in England, and studies on this topic gave inconsistent results(11, 17). Some were conducted on a local level(17) while others were conducted on a national level but used different methodologies and time periods(18). Therefore, research into the factors that affect the geographical variation of MMR vaccine uptake and coverage in England is needed to be able to tackle this problem by implementing targeted programs.

## 5. STUDY OBJECTIVES

The primary objective of the study is to determine whether there is a relationship between income deprivation measures in upper-tier local authorities and MMR vaccine uptake and coverage in England.

The secondary objective is to determine the effect of spatial clustering of income data on vaccination uptake and coverage.

## 6. STUDY TYPE

Geographical ecological study based on retrospective, publicly available data. The geographical units are upper-tier local authorities in England. STROBE 2007 (v4) Statement checklist for cross-sectional studies was used for the protocol design and is available in the S2 appendix.

## 7. TIMING OF FINAL ANALYSIS

Outcomes will be analysed collectively, as the data are retrospective and published online prior to the start of the analysis.

## 8. THE OUTCOME VARIABLES AND THEIR CALCULATION

The outcome variables are MMR vaccine uptake by 2 years of age (first dose) and coverage by 5 years of age (second dose) on upper-tier local authority geographical level. Both outcomes are continuous and represented as percentages. The whole population vaccination data will be obtained from national statistics which are routinely collected by Public Health England as part of the Cover of Vaccination Evaluated Rapidly (COVER). COVER extracts its data from Child Health Information Systems (CHIS) and from General Practices (GP) systems in a few local authorities(21), which is a good representation of the children population in England.

Vaccine coverage percentages were calculated before the data were publicly published by Public Health England. The calculation was conducted by dividing the number of eligible populations who were immunised in each upper-tier local authority (LA) by the number of eligible populations in that local authority, then multiplied by 100. The eligible population was defined as “the total number of children in the LA responsible population, reaching their nth birthday in the collection year”(21). The eligible population includes both, children registered with a GP in the local authority and those who are not registered but lived in that LA(21). The data is available for only 149 upper-tier local authorities, as 3 local authorities’ data were added to other LA(21). Data were published in September 2019(5) and covered the period from April 2018 to March 2019(22).

## 9. THE INDEPENDENT VARIABLES

Moran’s Index will be included in the analysis as an indicator for spatial clustering of income deprivation in upper-tier LA. The calculation methodology will be based on that adopted by Nyanzu and Rae (2019) using R software(23). It will involve income deprivation scores in Lower Layer Super Output Areas (LSOA) that constitute each local authority(23). Moran’s Index equation that will be used for the calculations is shown below:

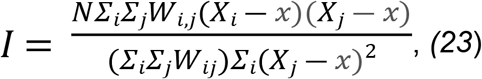

*Where*

*I* is the Global Moran’s Index;

*N* Is the number of observations.

*X*_*i*_ is the X variable when at area i.

*X*_*j*_ is the X variable when at area j.

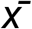 is the mean of the X variable.

*Wij* is the spatial weight used to compare area i and area j *(23*).

Deprivation gap data were published on a lower-tier local authority level, thus their average in each upper-tier local authority will be calculated. These variables were published by the Office for National Statistics and are based on data for the English Indices of Multiple Deprivation, 2019(24).

The income deprivation score, as well as the Income Deprivation Affecting Children Index (IDACI) 2019 on upper-tier local authority level will be included in the analysis. Data are available for 151 Upper-tier local authorities in England(25). A higher income deprivation score means that the area is more deprived(25). The 2019 Deprivation data collected represent the years 2015/2016 and are the latest published data(25).

Data are publicly available, free to use, published by the Ministry of Housing, Communities & Local Government and contain public sector information licensed under the Open Government Licence v3.0(26).

### 9.1 Covariates and confounding factors

To adjust for possible confounding factors, the following variables will be added to the multiple linear regression analysis. These are rural/urban classifications of upper-tier local authorities(27). This variable will be recoded using the dummy method to allow it to be included in the multiple linear regression.

Additionally, the live births rate for mothers’ age group in local authorities will be used as a proxy for the variation in mothers’ age in different local authorities. Data for the year 2016(28) will be used with the analysis of the uptake outcome variable, and data for the year 2013(29) will be used for the coverage outcome variable. The percentage of females aged 16-49 years in each category of highest qualification achieved (total 7 categories) in a LA will also be included in the analysis as a measure of variation in education across different local authorities(30). The total number of females aged 16-49 years who are residents in a local authority, based on the 2011 census will be used as a denominator to calculate the percentage in the corresponding LA(31).

## 10. STUDY POPULATION AND DATA COLLECTION

The study population is children living in England who were eligible for the MMR vaccine by their second and fifth birthday in 2018/2019 and who were registered with a GP in an upper-tier local authority or those who lived in the corresponding upper-tier local authority if they were not registered with a GP(22). Data will be collated from datasets published by the aforementioned sources and linked by upper-tier local authority’s codes and names in a new excel spreadsheet.

## 11. DATA ANALYSIS

### 11.1 Assumptions’ testing

Our initial choice of modelling the outcome variable will be to use a Linear Regression, hence we will test for the key assumptions of simple linear regression. The residuals of each outcome variable will be tested for normality visually by assessing a Q-Q plot and statistically by the Shapiro-Wilk test(32) at a significance level of 0.05, where the null hypothesis would be that the sample distribution is normal. If the result is significant, then the distribution will be regarded as non-normal.

The homoscedasticity assumption will also be tested before conducting the linear regression by plotting the standardised residuals against the standardised predicted values on a scatterplot. Moreover, the independent errors’ assumption will also be tested by plotting the standardized residuals against standardized predicted values. Randomly dispersed data with a rectangle shape and values lying between -3 and 3 on both the y and x-axes will be regarded as independent. Linearity will also be assessed for each independent variable versus each outcome by plotting them on a scatterplot.

We will consider other statistical approaches to transforming the outcome variable if we determine that it is not normally distributed for example by log-transformation. Alternatively, we would also consider adopting a count model, such as Poisson or Negative binomial regressions.

Finally, for multiple linear regression, Cook’s distance will be used to determine whether there are outliers that can influence the model and introduce bias, as well as the multicollinearity of independent variables through the Variance Inflation Factor (VIF) method and correlation coefficients.

### 11.2 Summary statistics

If the data are normally distributed, description of the location and dispersion of outcome variables will be described by mean, and Standard Deviation (SD). Otherwise, the median, and interquartile range will be described. Data will be represented using a box-and-whisker plot.

Percentages will be used for categorical data as a summary statistic and the number of final local authorities to be included will also be described.

### 11.3 Statistical tests

A two-sided significance level will be set at 0.05 for all relevant statistical tests.

Simple Linear Regression will be conducted after testing for its assumptions as previously mentioned.

A regression coefficient will be determined, as well as R ^2^ and adjusted P-value. A R^2^ > 70% will be considered to have a strong effect size, whereas, an R^2^ between 50% and 70% will be considered to have a moderate effect size, as adopted by Brennan, Moore and Millar (2022)(33).

Afterwards, multiple linear regression for the independent variables that were found to be associated with the outcome will be conducted after testing the relevant additional assumptions mentioned earlier. The standardized coefficients will be compared, in addition to the adjusted R^2^ and the P-value. Cook’s distance values greater than 1 will also be reported. Both statistical analyses will be conducted for both outcome variables.

Mallows Cp will be used to evaluate regression models. A value less than or equal to the number of parameters in any suggested model will be regarded as an unbiased model except for the full parameters model which will not be evaluated based on Mallow’s Cp. Bayesian Information Criterion (BIC) and Akaike Information Criterion (AIC) will also be used as complementary methods to Mallow’s Cp for the evaluation of regression models. The models will be compared and the ones with the lowest values for BIC and AIC will be shortlisted.

To analyse the secondary outcome, if the income deprivation score and Moran’s index are found to be associated with the outcomes, then mediation analysis will be conducted. Bootstrapping will be conducted afterwards to test for mediation effect significance.

An alternative method if the outcome variables’ distribution is non-normal is Generalized Linear Model (GLM). The independence of outcome variables’ observations will be checked before performing it, as well as determining the type of distribution and link function of the data.

## 12. HANDLING MISSING DATA

Six Local authorities will be excluded from the analysis. These are the City of London, Hackney, Rutland, Leicestershire, Cornwall, and the Isles of Scilly. This is because MMR coverage data for the City of London was reported under Hackney. Also, data for Rutland were reported under Leicestershire, in addition to Cornwall which contains data for the Isles of Scilly. The merge of two LA vaccination coverage data into one would affect the results of the analysis, taking into consideration the significant differences in income deprivation scores between them. For instance, the income deprivation rank of average score in the City of London is 144 and that of Hackney is 17, where 1 is the most deprived and 151 is the least deprived.

## 13. STATISTICAL PACKAGES

The data will be analysed using SPSS and Stata software, as well as Rstudio 2022.07.1+ 554.

## 14. STRENGTHS AND LIMITATIONS

To begin with, this study’s potential strengths, lie in its focus on tackling the re-emergence of MMR and trying to determine the factors that could have an impact on MMR uptake and coverage in England which in turn could have resulted in the removal of the elimination status of measles and rubella by the World Health Organization in 2019. It also aims to determine whether spatial clustering of income level influences the uptake and coverage of MMR vaccine in England.

On the other hand, a limitation of this work would be the use of aggregated data, meaning that further individual-level studies /research is required on this topic before being able to generalize the results.

## Data Availability

Collated research data will be made publicly available when the study is completed and published.

## 15. ETHICS

Ethical approval will not be required for this research because it will use publicly available data and does not seek to work with or identify personal identifiers.

## 16. DISSEMINATION

The results of the study would be discussed with key stakeholders through Personal and Public Involvement (PPI). This would be facilitated by the Hull York Medical School’s PPI coordinator. It is also intended to be disseminated through peer-reviewed journals and other media coverage.

## 17. AUTHORS CONTRIBUTIONS

Salma Ghazal: Conceptualization, Data Curation, Formal Analysis, Methodology, Writing – Original Draft Preparation, Writing – Review & Editing.

Ireneous Soyiri: Conceptualization, Formal Analysis, Supervision, Writing – Review & Editing.

## 18. Supporting Information

**S1 Appendix:** Data dictionary

| Variable | Variable name | Measurement unit | Allowed values | Variable definition | Description | References |
| --- | --- | --- | --- | --- | --- | --- |
| LA_code | Local authority code | ID String | E06000001 - E10000034 | Upper-tier local authority code. | A unique code assigned to each Upper-tier LA. | (25) |
| LA_name | Local authority name | ID String | text | Upper-tier local authority name. | A unique name given to each Upper-tier LA. | (25) |
| MMR_uptake | MMR vaccine uptake | Numeric Percentage | 74.3 to 97.1 % | The percentage of MMR vaccine uptake by 2 years of age (first dose) in upper-tier local authorities. | The number of eligible populations who were immunised by their second birthday in each upper-tier local authority (LA) divided by the number of eligible populations in that local authority x 100. | (5) |
| MMR_coverage | MMR vaccine coverage | Numeric Percentage | 64.1 to 96.4 % | The percentage of MMR vaccine coverage by 5 years of age (second dose) in upper-tier local authorities. | The number of eligible populations who were immunised with 2 doses of the MMR vaccine by their fifth birthday in each upper-tier local authority (LA) divided by the number of eligible populations in that local authority x 100. | (5) |
| MoransI | Moran's Index | Numeric Score | -1 to 1 | It is a measure of spatial autocorrelation, measuring the clustering of income deprivation. | 1 Perfect clustering<br>-1 Perfect dispersion<br>0 Randomness | (23) |
| Deprivation_gap | Deprivation gap | Numeric Percentage | 8.4 - 56.7% | It is the difference between the maximum 'Income Score' and the minimum 'Income Score' in each lower-tier local authority. | An average deprivation gap in upper-tier LA is calculated for this variable. | (24) |
| Income_deprivation_score | Income deprivation average score | Numeric Average score | 0.029 to 0.251 | A score based on combining the values returned from indicators related to income. | Income average score per upper-tier local authority. It is calculated by compiling data from LSOAs (population-weighted averages). Areas with the highest scores are the most deprived. | (25) |
| IDACI | Income Deprivation Affecting Children Index (IDACI) | Numeric Proportion | 0.032 to 0.327 | The proportion of children aged 0-15 years, who live in income-deprived families. | A proportion. | (25) |
| Rural_urban | Rural/urban classification of upper-tier local authorities | Categorical data (3 categories) | Predominantly Urban<br>Urban with Significant Rural<br>Predominantly Rural | The rural/urban classification of upper-tier local authorities is based on the percentage of the population living in rural areas. | The classification is based solely on the percentage of the population residing in rural areas in the specified geographical unit. Predominantly rural is where $\geq 50\%$ of the population lives in rural areas. Predominantly urban is where $\geq 74\%$ of the population reside in urban areas, whereas, urban with significantly rural is where 26 to 49% of the population live in rural areas. | (34) |
| Births_rate2016 | Live births rate per mothers' age group in 2016 | Numeric Rate | 0.3-171.3 | Live births rate per 1000 women per age group in every LA, in 2016. | Rate per 1000 women.<br>The age groups of mothers at birth are under 20, 20-24, 25-29, 30-34, 35-39, 40-44, 45 and over. | (28) |
| Births_rate2013 | Live births rate per mothers' | Numeric Rate | 0.2-169.9 | Live births rate per 1000 women per age | Rate per 1000 women.<br>The age groups of mothers at birth are under 20, 20- | (29) |

|  | age group in 2013 |  |  | group in every LA, in 2013. | 24, 25-29, 30-34, 35-39, 40-44, 45 and over. |  |
| --- | --- | --- | --- | --- | --- | --- |
| Education | The highest level of qualification achieved. | Numeric Percentage | 0.216 - 71.482% | The percentage of females aged 16 to 49 years per each (highest) level of qualification achieved in a local authority per total female population aged 16-49 years in that LA. | The levels of qualifications are:<br>• No qualifications;<br>• Level 1: 1-4 O Levels/CSE/GCSEs (any grades), Entry Level, Foundation Diploma, NVQ Level 1, Foundation GNVQ, Basic/Essential Skills;<br>• Level 2: 5+ O Level (Passes)/CSEs (Grade 1)/GCSEs (Grades A*-C), School Certificate, 1 A Level/ 2-3 AS Levels/VCEs, Intermediate/Higher Diploma, Welsh Baccalaureate Intermediate Diploma, NVQ level 2, Intermediate GNVQ, City and Guilds Craft, BTEC First/General Diploma, RSA Diploma;<br>• Apprenticeship;<br>• Level 3: 2+ A Levels/VCEs, 4+ AS Levels, Higher School Certificate, Progression/Advanced Diploma, Welsh Baccalaureate Advanced Diploma, NVQ Level 3; Advanced GNVQ, City and Guilds Advanced Craft, ONC, OND, BTEC National, RSA Advanced Diploma;<br>• Level 4 and above: Degree (for example BA, BSc), Higher Degree (for example MA, Ph.D., PGCE), NVQ Level 4-5, HNC, HND, RSA Higher Diploma, BTEC Higher level, Foundation degree (NI), Professional qualifications (for example teaching, nursing, accountancy);<br>• Other qualifications: Vocational/Work-related Qualifications, Foreign Qualifications (not stated/level unknown). | (30) |

**S2 Appendix:** STROBE checklist **STROBE 2007 (v4) Statement—Checklist of items that should be included in reports of *cross-sectional studies***

| Section/Topic | Item # | Recommendation | Reported on page # |
| --- | --- | --- | --- |
| Title and abstract | 1 | (a) Indicate the study's design with a commonly used term in the title or the abstract | p.1 and 2 |
|  |  | (b) Provide in the abstract an informative and balanced summary of what was done and what was found | p.2 |
| Introduction |  |  |  |
| Background/rationale | 2 | Explain the scientific background and rationale for the investigation being reported | p.3 |
| Objectives | 3 | State specific objectives, including any prespecified hypotheses | p.4 |
| Methods |  |  |  |
| Study design | 4 | Present key elements of study design early in the paper | p.4 |
| Setting | 5 | Describe the setting, locations, and relevant dates, including periods of recruitment, exposure, follow-up, and data collection | p.4 and 5 |
| Participants | 6 | (a) Give the eligibility criteria, and the sources and methods of selection of participants | NA |
| Variables | 7 | Clearly define all outcomes, exposures, predictors, potential confounders, and effect modifiers. Give diagnostic criteria, if applicable | p. 4 and 5 |
| Data sources/ measurement | 8* | For each variable of interest, give sources of data and details of methods of assessment (measurement). Describe comparability of assessment methods if there is more than one group | p. 4 and 5 |
| Bias | 9 | Describe any efforts to address potential sources of bias | p.5 |
| Study size | 10 | Explain how the study size was arrived at | NA |
| Quantitative variables | 11 | Explain how quantitative variables were handled in the analyses. If applicable, describe which groupings were chosen and why |  |
| Statistical methods | 12 | (a) Describe all statistical methods, including those used to control for confounding | p.5, 6 and 7 |
|  |  | (b) Describe any methods used to examine subgroups and interactions | NA |
|  |  | (c) Explain how missing data were addressed | p.7 |
|  |  | (d) If applicable, describe analytical methods taking account of sampling strategy | NA |
|  |  | (e) Describe any sensitivity analyses | NA |
| Results |  |  |  |
| Participants | 13* | (a) Report numbers of individuals at each stage of study—eg numbers potentially eligible, examined for eligibility, confirmed eligible, included in the study, completing follow-up, and analysed | NA |
|  |  | (b) Give reasons for non-participation at each stage | NA |
|  |  | (c) Consider use of a flow diagram | NA |
| Descriptive data | 14* | (a) Give characteristics of study participants (eg demographic, clinical, social) and information on exposures and potential confounders | p.9 |
|  |  | (b) Indicate number of participants with missing data for each variable of interest | p.7 |
| Outcome data | 15* | Report numbers of outcome events or summary measures | p.4 |
| Main results | 16 | (a) Give unadjusted estimates and, if applicable, confounder-adjusted estimates and their precision (eg, 95% confidence interval). Make clear which confounders were adjusted for and why they were included | NA |
|  |  | (b) Report category boundaries when continuous variables were categorized | NA |
|  |  | (c) If relevant, consider translating estimates of relative risk into absolute risk for a meaningful time period | NA |
| Other analyses | 17 | Report other analyses done—eg analyses of subgroups and interactions, and sensitivity analyses | NA |
| Discussion |  |  |  |
| Key results | 18 | Summarise key results with reference to study objectives | NA |
| Limitations | 19 | Discuss limitations of the study, taking into account sources of potential bias or imprecision. Discuss both direction and magnitude of any potential bias | p.7 and 8 |
| Interpretation | 20 | Give a cautious overall interpretation of results considering objectives, limitations, multiplicity of analyses, results from similar studies, and other relevant evidence | NA |
| Generalisability | 21 | Discuss the generalisability (external validity) of the study results | NA |
| Other information |  |  |  |
| Funding | 22 | Give the source of funding and the role of the funders for the present study and, if applicable, for the original study on which the present article is based | p.1 |

## REFERENCES

1. UK Health Security Agency. Measles: the green book, chapter 21: GOV.UK; 2019 [

2. Mulchandani R, Sibal B, Phillips A, Suleman S, Banerjee A, Teagle R, et al. A large outbreak of measles in the West Midlands, England, 2017-2018: descriptive epidemiology, control measures and lessons learnt. Epidemiology & Infection. 2021;149(e114):1–6.

3. Pegorie M, Shankar K, Welfare WS, Wilson RW, Khiroya C, Munslow G, et al. Measles outbreak in Greater Manchester, England, October 2012 to September 2013: epidemiology and control. Eurosurveillance. 2014;19(49).

4. Public Health England. Laboratory confirmed cases of measles, rubella and mumps, England: October to December 2019.; 2020.

5. NHS Digital. Childhood Vaccination Coverage Statistics - England 2018-19-NHS Digital.: NHS Digital; 2019 [Available from: https://digital.nhs.uk/data-and-information/publications/statistical/nhs-immunisation-statistics/england-2018-19.

6. NHS Digital. Childhood Vaccination Coverage Statistics- 2020-21.: NHS Digital; 2021 [Available from: https://digital.nhs.uk/data-and-information/publications/statistical/nhs-immunisation-statistics/england2020-21#summary.

7. Hungerford D, Macpherson P, Farmer S, Ghebrehewet S, Seddon D, Vivancos R, et al. Effect of socioeconomic deprivation on uptake of measles, mumps and rubella vaccination in Liverpool, UK over 16 years: a longitudinal ecological study. Epidemiology and Infection. 2016;144(6).

8. Baker D, Garrow A, Shiels C. Inequalities in immunisation and breast feeding in an ethnically diverse urban area: cross-sectional study in Manchester, UK. Epidemiology and Community Health. 2011;65(4):346–52.

9. Sandford H, Tata LJ, Browne I, Pritchard C. Is there an association between the coverage of immunisation boosters by the age of 5 and deprivation? An ecological study. Vaccine. 2015;33(9):1218–22.

10. Osam CS, Pierce M, Hope H, Ashcroft DM, Abel KM. The influence of maternal mental illness on vaccination uptake in children: a UK population-based cohort study. European Journal of Epidemiology. 2020;35(9):879–89.

11. Pearce A, Law C, Elliman D, Cole TJ, Bedford H. Factors associated with uptake of measles, mumps, and rubella vaccine (MMR) and use of single antigen vaccines in a contemporary UK cohort: prospective cohort study. BMJ. 2008;336(7647):754–7.

12. Torracinta L, Tanner R, Vanderslott S. MMR Vaccine Attitude and Uptake Research in the United Kingdom: A Critical Review. Vaccines. 2021;9(402).

13. Smith D, Newton P. Structural barriers to measles, mumps and rubella (MMR) immunisation uptake in Gypsy, Roma and Traveller communities in the United Kingdom. Critical Public Health. 2016;27(2):238–47.

14. Brown KF, Long SJ, Ramsay M, Hudson MJ, Green J, Vincent CA, et al. UK parents’ decision-making about measles–mumps–rubella (MMR) vaccine 10 years after the MMR-autism controversy: A qualitative analysis. Vaccine. 2012;30(10):1855–64.

15. Thompson EL, Rosen BL, Maness SB. Social Determinants of Health and Human Papillomavirus Vaccination Among Young Adults, National Health Interview Survey 2016. Journal of Community Health. 2018;44(2019):149–58.

16. Danis K, Georgakopoulou T, Stavrou T, Laggas D, Panagiotopoulos T. Socioeconomic factors play a more important role in childhood vaccination coverage than parental perceptions: a cross-sectional study in Greece. Vaccine. 2010;28(7):1861–9.

17. Hungerford D, Macpherson P, Farmer S, Ghebrehewet S, Seddon D, Vivancos R, et al. Effect of socioeconomic deprivation on uptake of measles, mumps and rubella vaccination in Liverpool, UK over 16 years: a longitudinal ecological study. Epidemiology & Infection. 2016;144(6):1201–11.

18. Wright JA, Polack C. Understanding variation in measles-mumps-rubella immunization coverage--a population-based study. European Journal of Public Health. 2006;16(2):137–42.

19. Ferguson KD, McCann M, Katikireddi SV, Thomson H, Green MJ, Smith DJ, et al. Evidence synthesis for constructing directed acyclic graphs (ESC-DAGs): a novel and systematic method for building directed acyclic graphs. International Journal of Epidemiology. 2020:322–9.

20. Textor J, Zander Bvd, Gilthorpe MK, Liskiewicz M, Ellison GTH. DAGitty — draw and analyze causal diagrams.: DAGitty.net; 2016 [Available from: http://www.dagitty.net.

21. Screening & Immunisations Team (NHS Digital), COVER Team (Public Health England). Childhood Vaccination Coverage Statistics Quality Statement: NHS digital; 2019 [

22. Public Health England. COVER programme: guide to submitting data v02.00. Public Health England; 2020.

23. Nyanzu E, Rae A. An English Atlas of Inequality - Technical Report.: GitHub; 2019 [Available from: http://alasdairrae.github.io/atlasofinequality/reports/tech_report_aoi_21_nov_2019.pdf.

24. Office for National Statistics. Mapping income deprivation at a local authority level.: Office for National Statistics; 2021 [Available from: https://www.ons.gov.uk/peoplepopulationandcommunity/personalandhouseholdfinances/incomeandwealth/datasets/mappingincomedeprivationatalocalauthoritylevel.

25. Ministry of Housing-Communities and Local Government. National statistics, English indices of deprivation 2019-File:11upper-tier local authority summaries.: GOV.UK; 2019 [Available from: https://www.gov.uk/government/statistics/english-indices-of-deprivation-2019.

26. McLennan D, Noble S, Noble M, Plunkett E, Wright G, Gutacker N. The English Indices of Deprivation 2019-Technical Report. Ministry of Housing-Communities & Local Government.; 2019.

27. Department for Environment-Food & Rural Affairs. Official Statistics 2011 Rural Urban Classification lookup tables for all geographies.: GOV.UK; 2021 [Available from: https://www.gov.uk/government/statistics/2011-rural-urban-classification-lookup-tables-for-all-geographies.

28. Haines N. Births by mothers’ usual area of residence in the UK.: Office for National Statistics (ONS); 2016 [Available from: https://www.ons.gov.uk/peoplepopulationandcommunity/birthsdeathsandmarriages/livebirths/datasets/birthsbyareaofusualresidenceofmotheruk.

29. Haines N. Births by mothers’ usual area of residence in the UK.: Office for National Statistics (ONS); 2013 [Available from: https://www.ons.gov.uk/peoplepopulationandcommunity/birthsdeathsandmarriages/livebirths/datasets/birthsbyareaofusualresidenceofmotheruk.

30. Statistics. N-OLM. Highest level of qualification by sex by age [DC5102EW] - 2011: NOMIS; 2013 [Available from: https://www.nomisweb.co.uk/census/2011/dc5102ew.

31. Office for National Statistics. Table PP03UK: Female usual resident population by single year of age, unrounded estimates, local authorities in the United Kingdom (Excel sheet 806Kb): Office for National Statistics.; 2013 [Available from: https://www.ons.gov.uk/peoplepopulationandcommunity/populationandmigration/populationestimates/datasets/2011censuspopulationestimatesbysingleyearofageandsexforlocalauthoritiesintheunitedkingdom.

32. Ghasemi A, Zahediasl S. Normality Tests for Statistical Analysis: A Guide for Non-Statisticians. Int J Endocrinol Metab. 2012;10(2):486–9.

33. Brennan OC, Moore JE, Millar BC. Does social deprivation correlate with meningococcal MenACWY, Hib/MenC and 4CMenB/Meningococcal Group B vaccine uptake in Northern Ireland? Ulster Medical Journal. 2022;91(1):9–18.

34. Defra Rural Statistics. The 2011 Rural-Urban Classification for Local Authority Districts in England: GOV.UK; 2017 [Available from: https://assets.publishing.service.gov.uk/government/uploads/system/uploads/attachment_data/file/591464/RUCLAD_leaflet_Jan2017.pdf.

